# A 6-week randomized-controlled field study: Effect of isokinetic eccentric resistance training on strength, flexibility and muscle structure for the shoulder external rotator in male junior handball players

**DOI:** 10.1101/2023.12.06.23299595

**Authors:** Sebastian Vetter, Maren Witt, Pierre Hepp, Axel Schleichardt, Stefan Schleifenbaum, Christian Roth, Timm Denecke, Jeanette Henkelmann, Hans-Peter Köhler

## Abstract

**Background:** Team handball involves a tremendous amount of shoulder motion with high forces during repeated extended external range of motion. Shoulder complaints are a common problem in youth handball players. While eccentric training for the lower extremity shows preventive effects by improving strength, range of motion and fascicle length, there is a research gap for the shoulder joint and for advanced tissue characterization using diffusion tensor imaging.

**Objectives:** To investigate the effects of six-week eccentric isokinetic resistance training on the strength, flexibility, and fiber architecture characteristics of the external rotators compared to an active control group in junior handball players.

**Methods:** 15 subjects were randomly assigned to the eccentric training group and 14 subjects to the active control group (conventional training). Primary outcome measures were eccentric and concentric isokinetic strength of the external rotators, range of motion, and muscle fascicle length and fascicle volume.

**Results:** The intervention group, showed significant changes in eccentric strength (+15 %). The supraspinatus and infraspinatus muscles showed significant increases in fascicle length (+13 % and +8 %), and in fractional anisotropy (+9 % and 6 %), which were significantly different from the control group.

**Conclusions:** Eccentric isokinetic training has a significant effect on the function and macroscopic structure of the shoulder external rotators in male junior handball players. While strength parameters and muscle structure improved, range of motion did not change. Future research is needed for more convenient field exercises in this context.

## 1. Introduction

The frequency and severity of shoulder injuries is an ongoing problem in overhead sports as the shoulder is the fourth most commonly injured joint at 9.1%.^1^ The prevalence of shoulder injuries reaches up to 36 %, and in baseball the return to performance rate is dramatically low on 7 %.^2^ As an example for early career deficits in shoulder, 69 % of junior javelin throwers revealed posterosuperior intraosseous cysts of the humeral head with a size larger than > 3 mm in their throwing shoulder compared to 15 % in the contralateral shoulder. Even when age cannot be found as a risk factor for injury, most future problems in shoulder can be addressed to chronic overuse beginning in early high performance sports career stage.^3,4^

A major reason for overuse injuries are the overhead demands due to the repetitive throwing motions. While most research exist for baseball pitching, showing high maximum internal rotation torque and high external range of motion (ROM) before ball release,^5^ also the deceleration phase puts high loads on the stabilizing shoulder tissues.^6^ Therefore, a throwing performance is always associated with high eccentric load at the very end of the individual ROM.^6^

In Handball similar demands^7^ and shoulder adaptations occur.^8^ Chronic discomfort, loss of shoulder performance or even acute injuries are mostly found with internal impingement and rotator cuff tears.^9^ In particular, 25% of youth handball players suffer overuse injuries to the shoulder each season, requiring longer rehabilitation than other joints.^10^ To primary prevent shoulder problems, there are several options. A typical approach is to improve flexibility and strength, which are considered risk factors for injury.^11^ A glenohumeral internal rotation deficit (GIRD) above 20° is typically found for a throwing shoulder with high risk for injury.^9^ Also, an isometric strength ratio of 0.78 for the external rotators was associated with overused and injured shoulders in youth male handball players.^4^ However, in handball, there is a gap in prevention studies for the shoulder.^4^ But prevention studies are desperately needed because common prevention programs do not reduce the risk of overuse injuries in handball.^12^ To the best of our knowledge, only Andersson et al.^13^ investigated the effects of a strengthening program for the external rotators and found an increase in strength and a 28% decrease in shoulder injuries, providing a little indication of the potential for shoulder prevention exercises.

To increase the impact of training interventions by addressing multiple risk factors at the same time, innovative approaches to strength training for the lower limb use eccentric-focused protocols. Recent review articles show that not only flexibility^14^ but also strength^15^ can be increased more advanced by eccentric-only exercises compared to concentric-focused exercises or passive stretching. In reality, even low level standardized field eccentric exercises implemented in a regular athletic training routine halved the risk of hamstring injuries, according to a recent meta-analysis of 8459 athletes.^16^ This effectiveness may be explained with high impact changes on macroscopic scale. Eccentric training produces tissue damage^17^ that promote hypertrophic reactions in terms of increasing muscle cross-sectional-area and muscle volume.^18^ Interestingly, besides muscle volume changes, a FL lengthening^17^ occurs after eccentric strength training compared to concentric strength training interventions^19^ or stretching.^20^ Since a shortened muscle fiber has also been suggested as a risk factor for lower extremity injury,^21^ exercises that promote fascicle lengthening may be a beneficial intervention for the shoulder as well.^22^

Especially in the last decade, an increasing number of highly standardized eccentric training studies have been published for the lower limbs, but not for the shoulder.^19^ Therefore, a research gap exists for eccentric strength training on the shoulder rotator cuff muscles^23^ and for healthy junior handball players as a sample with high shoulder muscle demands and complaints.^24^ Further, a fundamental research gap exists for the investigation of three-dimensional changes in muscle architecture.^25^ For this reason, quantitative magnetic resonance imaging (MRI) techniques are increasingly being used for technologically advanced tissue characterization. MRI-based diffusion tensor imaging of muscle (mDTI) is an innovative approach for showing the fiber arrangement for the whole muscle volume.^26^ This technique provides a valid,^27^ and reliable^28,29^ calculation of muscle fiber metrics and may solve problems existing in the use of 2D ultrasound imaging.

To address the aforementioned research gaps and issues, the aim of this study was to investigate the structural and functional changes of the dominant throwing shoulder in competitive male junior handball players after six weeks of isokinetic eccentric strength training compared to an active control group. Changes will be determined based on isokinetic dynamometry and mDTI. Primary outcome measures were the external rotator muscles active and passive ROM, maximal eccentric and concentric strength at 30°/s and 60°/s isokinetic velocity, the muscle fascicle length (FL), absolute fascicle volume (FV), and fractional anisotropy (FA) for the supraspinatus and infraspinatus muscle. Secondary outcome measures were the strength and flexibility parameters for the antagonistic internal rotator muscles.

## 2. Materials and methods

This study is a randomized-controlled trial with an exercise-based intervention. Five days before (pre) and after (post) the training phase, functional and structural parameters were examined for the dominant throwing shoulder using mDTI and an isokinetic dynamometer as described below. The participants were recruited locally in the pre-season of the German national junior league in August 2022. The study can be found in the German Register for Clinical Trials (DRKS00028885). The study protocol was approved by the Leipzig University ethics committee (ethical approval number: 362/21-ek). The study was conducted in accordance with the relevant guidelines and regulations. Written informed consent was obtained from all subjects.

### 2.1 Sample

The sample consisted of 29 male junior handball players (German male handball division A and U23) from a local elite handball club (18.0 ± 1.6 years; 186.8 ± 6.3 cm; 84.8 ± 11.3 kg). Sample size was based on a preliminary study^30^ and on comparable studies that calculated or suggested 13 to 16 participants per group.^23,31^ The participants were randomly assigned to one of the two groups according to their playing position. Participants had to show a healthy throwing shoulder. Therefore, exclusion criteria were regular medication that may affect the adaptability, muscle-nerve diseases, discomfort, pain or known lesions of the right shoulder. Further, subjects were told to stop with unregular physical activity or intense training five days prior to data collection.

### 2.2 Intervention group eccentric exercise

Altogether, 12 eccentric isokinetic strength training sessions for the external rotators were performed within six weeks (twice weekly). The training was performed on a BTE Primus RS isokinetic dynamometer (Baltimore Therapeutic Equipment Company, Hanover, MD, USA). All subjects completed two familiarization sessions before intervention phase. The eccentric isokinetic training was incorporated into the regular athletic training routine, but replaced specific preventive shoulder exercises performed instead by the active control group. The eccentric training protocol was standardized as follows: 5×10 repetitions with 90 seconds rest in between, ±10 % load magnitude based on sets, four seconds of time under tension per repetition, 120° ROM, 30°/s movement velocity, supine position with 90° elbow flexion and neutral hand position, 48-76 hours rest between training sessions. After three weeks of intervention, the total ROM was shifted approximately 15° toward the internal ROM and the load magnitude was increased based on intensity of the previous three sessions.

### 2.3 Control Group conventional training

The active control group performed common preventive exercises in athletic training as recommended by the German Handball Federation in the athletics concept.^32^ Depending on the needs of the local handball club, the prevention exercises included different free-weight rotator cuff exercises such as shoulder abduction, front rowing, and shoulder rotation. The training load was adapted and equalized to the eccentric training group in terms of weeks and days of training, and sets and repetitions in each session.

### 2.4 Isokinetic Testing

Flexibility and strength tests were performed prior to MRI on the IsoMed 2000 isokinetic dynamometer (D&R Ferstl GmbH, Hemau, Germany). As described for eccentric training, subjects were tested for the dominant throwing shoulder in the supine position with fixed shoulder-arm joints (90° shoulder abduction, 90° elbow flexion). To prevent shoulder elevation, the shoulder was also fixed ventrally. In terms of order, a 10-minute warm-up on the rowing machine was performed first. Then, the isokinetic diagnostics began with active and passive stretching tests, followed by strength tests at 60°/s and then at 30°/s isokinetic velocity, always separated for concentric and eccentric movement. About 1 minute of rest was allowed between tests.

The active stretch test consisted of two familiarization repetitions and five consecutive test trials that dynamically alternated between internal and external rotation direction. When the subject was unable to continue the antagonist stretch produced by the agonist muscles, the maximum active ROM and the turning point were reached. Passive tests were performed at an isokinetic speed of 10°/s. The passive stretch consisted of ten trials of alternating internal and external rotation with the eyes closed. Maximum ROM was reached when subjects either reached 9 Nm or 100° of internal rotation and 140° of external rotation.

The maximal strength tests were performed separately for concentric and eccentric, also alternating between internal and external rotation. After two familiarization trials, three repetitions were performed over an amplitude of 150° (60° internal rotation and 90° external rotation) at an isokinetic velocity of 60°/s and then 30°/s. All tests were conducted in the same order. The position of each subject, the dynamometer arm and gravity correction as well as the settings were individualized at baseline and applied in the same way for the post testing. The data were recorded on the isokinetic dynamometer with a recording frequency of 200 Hz.

### 2.5 Diffusion Tensor Imaging

In supine position MRI scans were performed using a 3-Tesla Siemens MAGNETOM Prisma scanner (Siemens Healthcare, Erlangen, Germany) and a shoulder coil (XL, 16-channel). Commercial T1-weighted (T1w) and diffusion-weighted imaging sequences (DWI) were used. The total scan time was twelve minutes. The volunteers lay in a head first supine position with the right arm in the neutral position and the hand supinated. For the T1w the following settings were acquired: TR/TE = 492/20 ms, slice thickness = 0.7 mm, flip angle = 120°, FOV = 180 x 180 mm^2^, matrix = 256×256 mm^2^. The following 2D echo planar DWI sequence was acquired as follows: TR/TE = 6100/69 ms, slice thickness = 4 mm, flip angle = 90°, FOV = 240 x 240 mm^2^, matrix = 122 x 122 mm^2^, 48 diffusion sampling directions with b = 400 s/mm^2^.

### 2.6 Data processing

The isokinetic dynamometer data were processed using MATLAB v. R2022a (MathWorks, Natick, USA). Raw-data were filtered using a fifth order, zero-lag Butterworth low-pass filter at a cut-off frequency of 6 Hz. Afterwards, the torque and angle data were cut according to repetition number and movement direction (internal or external rotation). From each repetition the acceleration and deceleration phase were cut. The maximum and average torque were calculated from the resulting torque-angle curves for each movement direction (internal/ external rotation) and contraction mode (eccentric/ concentric) per participant. For further analysis, the torque and angle data were normalized to body mass. In order to compare torque-angle-curves, the data were interpolated to 101 samples for one-dimensional Statistical Parametric Mapping (SPM1d).^33^ Flexibility analysis were based on the average of the five trials of each active and passive stretching test. For these trials, a three-parametric e-function was fitted.^30^ This enabled analyses for submaximal ROM and maximal ROM. SPM1d was also used to analyze the passive torque-angle curves.

MRI data were processed using Mimics Materialise v. 24.0 (Leuven, Belgium) for prior muscle segmentation and DSI Studio (v. 3 December 2021, http://dsi-studio.labsolver.org) for muscle tractography. The T1w images from the MRI were used for manual segmentation of the supraspinatus and infraspinatus muscles. After extraction of the segmented muscle volume of interest, the DWI were corrected for motion and eddy current distortion using DSI Studio’s integrated FSL eddy current correction. The DWI were visually inspected by two independent raters. If artifacts or field of view did not allow full muscle analysis, subjects were excluded. Tractography was performed using specific stopping criteria defined in previous work.^29^ The deterministic fiber tracking is based on an RK4 algorithm. After tractography, DSI Studio was used to calculate the muscle FL and the FV using the integrated statistics tool. This method has been previously evaluated.^29^

### 2.7 Statistics

Statistics and graphs were performed using MATLAB v. R2022a (MathWorks, Natick, USA) and SPSS v. 27 (IBM, Armonk, New York, USA). Descriptive results were based on mean values and the standard deviation (±). Participants were excluded from further analysis if the z-transformed values reached 2.5. Repeated measures mixed multivariate analysis of variance (MANOVA) were used to show overall differences. The factors were group (Intervention and control), time (pre and post), mode (eccentric and concentric), and speed (30°/s and 60°/s). Post-hoc analysis was performed using repeated measures univariate analysis of variance (ANOVA) and t-tests for within group pre-post comparisons. The level of significance was set to α = 0.05. Further, to compare the morphology of the angle-torque and passive torque-angle trajectories, the interpolated curves were compared using SPM1d. For this purpose, a two-factor (time and group) ANOVA with repeated measures was chosen.

## 3. Results

14 subjects from the intervention group (18.0 ± 1.1 years; 188.3 ± 6.8 cm; 88.9 ± 11.6 kg) and 13 subjects from the active control group (17.9 ± 2.2 years; 184.1 ± 5.1 cm; 79.2 ± 9.4 kg) could be included in analysis. These subjects completed 12 training sessions within six weeks with sufficient compliance. However, from this sample, 22 participants were eligible for supraspinatus fascicle analyses due to limited field of view and 25 participants were included in the isokinetic data analyses, as four participants were not available for functional testing due to shoulder problems or transfer to another handball club.

### 3.1 Primary outcome measures

Functional and structural changes for the trained dominant throwing shoulder external rotator cuff muscles were analyzed for pre-post changes within the intervention and control group and for differences in changes between the two groups as shown in Table 1.

**Table 1:**
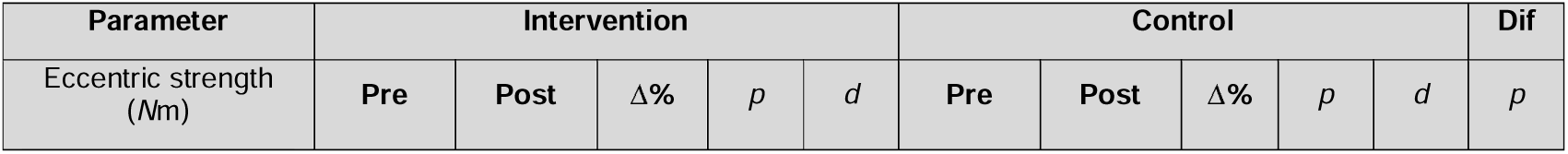
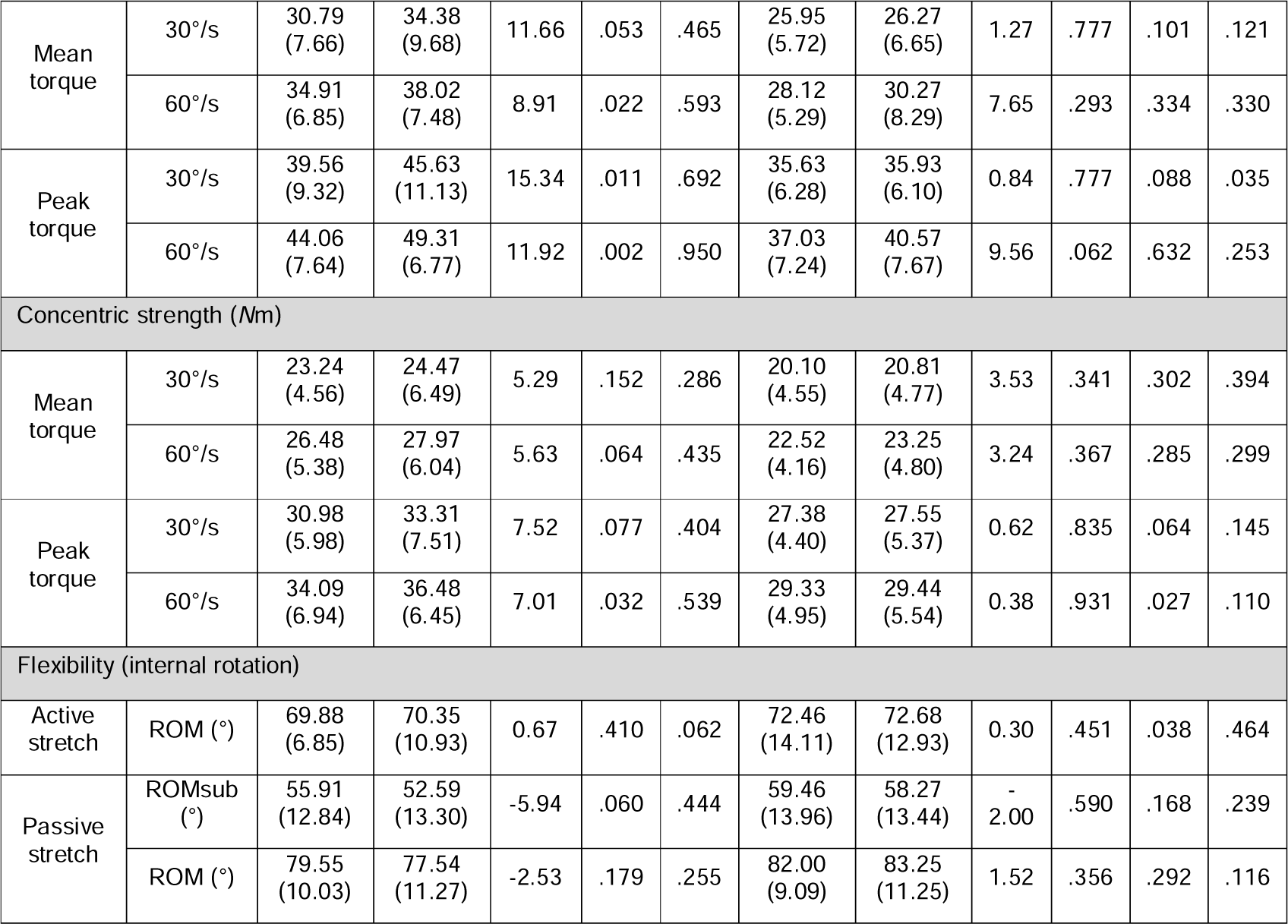
Primary outcome measures post-hoc comparisons. Columns indicate the absolute pre and post-test values and in brackets the values for standard deviation for each group, percentage change (Δ%) and significant pre-post differences within groups (*p*), cohens d effect size (*d*), and in column “Dif” the statistical significance for between-group differences (normalized to body mass for strength). Nm, newton meter; ROMsub, submaximal range of motion; °, angle degree; °/s, degrees per second (movement velocity).

For strength measures (normalized to body mass), an overall four-way mixed measures MANOVA for group x time x mode x speed showed a main effect for the factor time (Wilks-Lambda = .699; F(2,22) = 4.74; *p* = .019; ηp^2^ = . 301), an interaction effect for time x mode (Wilks-Lambda = .699; F(2,22) = 4.74; *p* = .019, ηp^2^ = .301) and an interaction effect for the factors time x speed (Wilks-Lambda = .691, F(2,22) = 4.92; *p* = .017; ηp^2^ = .309) but not for time x group (Wilks-Lambda = .832; F(2,22) = 4.74; *p* = .132; ηp^2^ = .168). Post-hoc analysis revealed a group x time interaction in peak torque and thus a training effect for the trained 30°/s eccentric strength (F(1,23) = 3.628, *p* = .035; ηp^2^ = .136). The main effect time showed group independent significant differences from pre to post (F(1,23) = 4.82, *p* = .038, ηp^2^ = .173).

For flexibility measures, a two-way MANOVA (group x time) revealed no interaction effect for the passive stretching test for the external rotators (Wilks-Lambda = .945; F(2,22) = <1; *p* = .054; ηp^2^ = .055). Furthermore, the active stretching revealed no significant interaction effect for the factors group x time (F(1,23) = <1; *p* = .773; ηp^2^ = .004). Further post-hoc comparisons for strength and flexibility parameters can be found in Table 1.

For analysis of the supraspinatus muscle fascicle, a two-way MANOVA showed a group x time-interaction effect (Wilks-Lambda = .492; F(3,17) = 5.86; *p* = .006; ηp^2^ = .508). Furthermore, ANOVA showed a significant change in fascicle length (F(1;19) = 3.37; *p* = .041; ηp^2^ = .151) and FA (F(1,19) = 3.92; *p* = .062; ηp^2^ = .171). The infraspinatus muscle also showed positive interaction effects for group x time (Wilks-Lambda = .281; F(3,11) = 9.38; *p* = .002; ηp^2^ = .719). The univariate analysis revealed a positive change for fascicle length (F(1,13) = 4.17; *p* = .031; ηp^2^ = .243) and FA (F(1,13) = 10.25; *p* = .007; ηp^2^ = .441). Further post-hoc comparisons can be found in Table 2.

**Table 2:** Post-hoc results for muscle structural measures. Columns indicate the absolute pre and post-test values and in brackets the values for standard deviation for each group, percentage change (Δ%) and significant pre-post differences within groups (*p*), cohens d effect size (*d*), and in column “Dif” the statistical significance for between-group differences.

| Muscle | parameter | Intervention | | | | | Control | | | | | Dif<br>$p$ |
| --- | --- | --- | --- | --- | --- | --- | --- | --- | --- | --- | --- | --- |
| | | Pre | Post | $\Delta\%$ | $p$ | $d$ | Pre | Post | $\Delta\%$ | $p$ | $d$ | |
| Infraspinatus | Fascicle length (mm) | 69.47<br>(13.31) | 75.23<br>(7.82) | 8.29 | .038 | .632 | 81.04<br>(7.98) | 77.35<br>(6.82) | -4.55 | .287 | .549 | .031 |
|  | Fascicle volume | 273.78 | 291.49 | 6.47 | .248 | .224 | 305.05 | 297.60 | -2.44 | .749 | .154 | .265 |
|  | (mm<sup>3</sup>) | (79.84) | (48.58) |  |  |  | (61.38) | (19.73) |  |  |  |  |
|  | Fractional Anisotropy | .2510<br>(.0274) | .2732<br>(.0243) | 8.85 | .002 | 1.251 | .2792<br>(.0196) | .2671<br>(.0141) | -4.33 | .307 | .523 | .004 |
| Supraspinatus | Fascicle length (mm) | 37.47<br>(6.42) | 42.38<br>(6.17) | 13.10 | .017 | .663 | 39.99<br>(8.00) | 39.59<br>(6.45) | -1.00 | .819 | .079 | .041 |
|  | Fascicle volume (mm<sup>3</sup>) | 153.18<br>(37.29) | 165.65<br>(26.52) | 8.14 | .158 | .303 | 146.65<br>(41.34) | 164.07<br>(22.37) | 11.88 | .246 | .418 | .395 |
|  | Fractional Anisotropy | .2849<br>(.0267) | .3029<br>(.0141) | 6.32 | .028 | .584 | .2947<br>(.0266) | .2891<br>(.0281) | -1.90 | .386 | .306 | .031 |

For the external rotator muscles eccentric strength, SPM1d analysis showed a significant group x time difference and an increase in the torque-angle relationship in between 30 and 60° internal ROM (Figure 1). The passive torque-angle curves showed an increase in passive stiffness for the internal rotation direction in the intervention group, but neither internal nor external rotation movements showed statistically significant pre-post changes (Figure 2).

**Figure 1:**
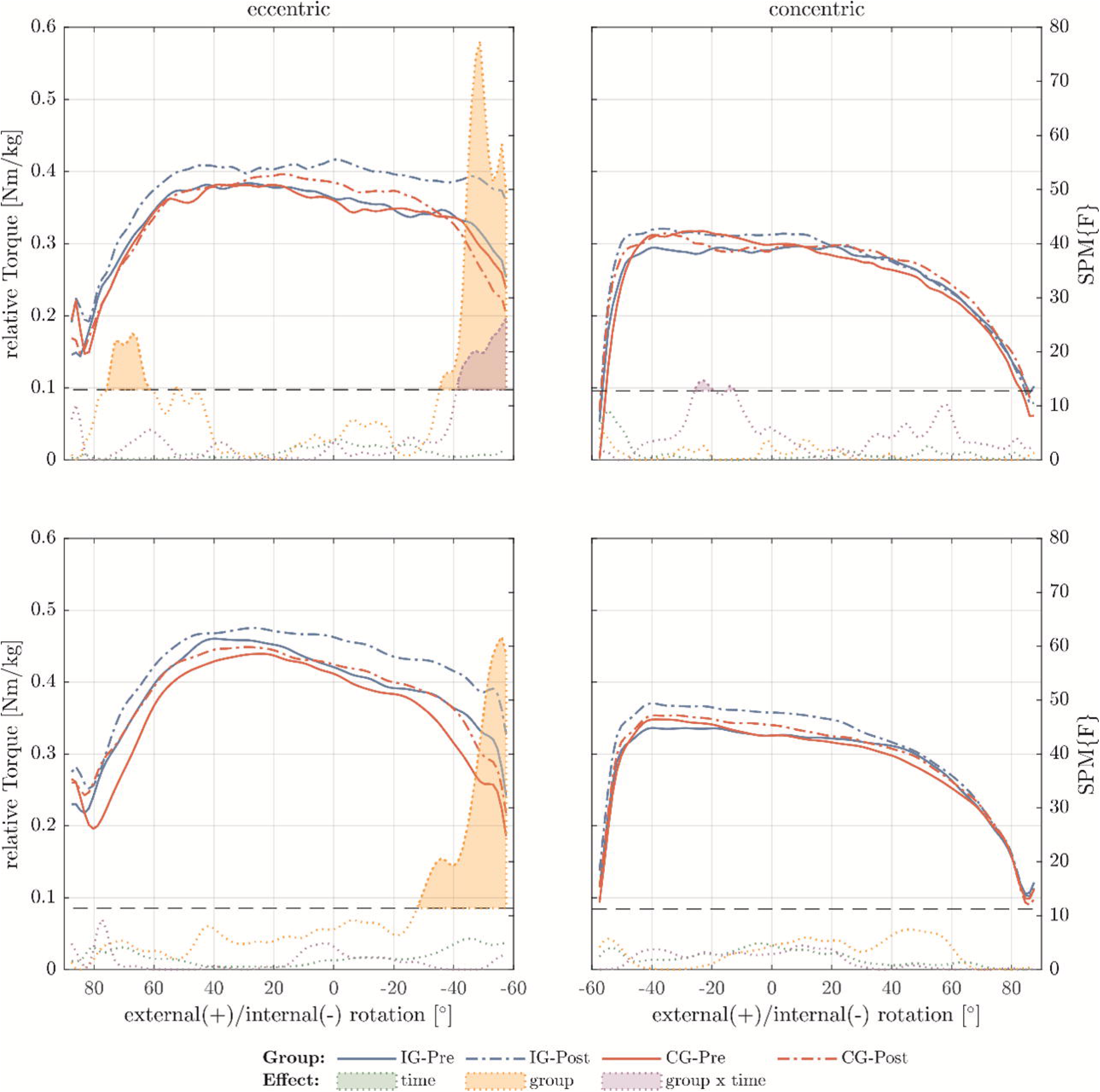
Relative torque-angle curves for 30°/s (top row) and 60°/s (bottom row) of the external rotators for the intervention group (IG) and control group (CG) at the different time points of pre- and post-test. Additionally the F-value functions for the different effects time, group and time x group calculated using SPM1d. Values exceeding the critical F-value (dashed horizontal line) show significant differences for the different effects and are filled with the corresponding color

**Figure 2:**
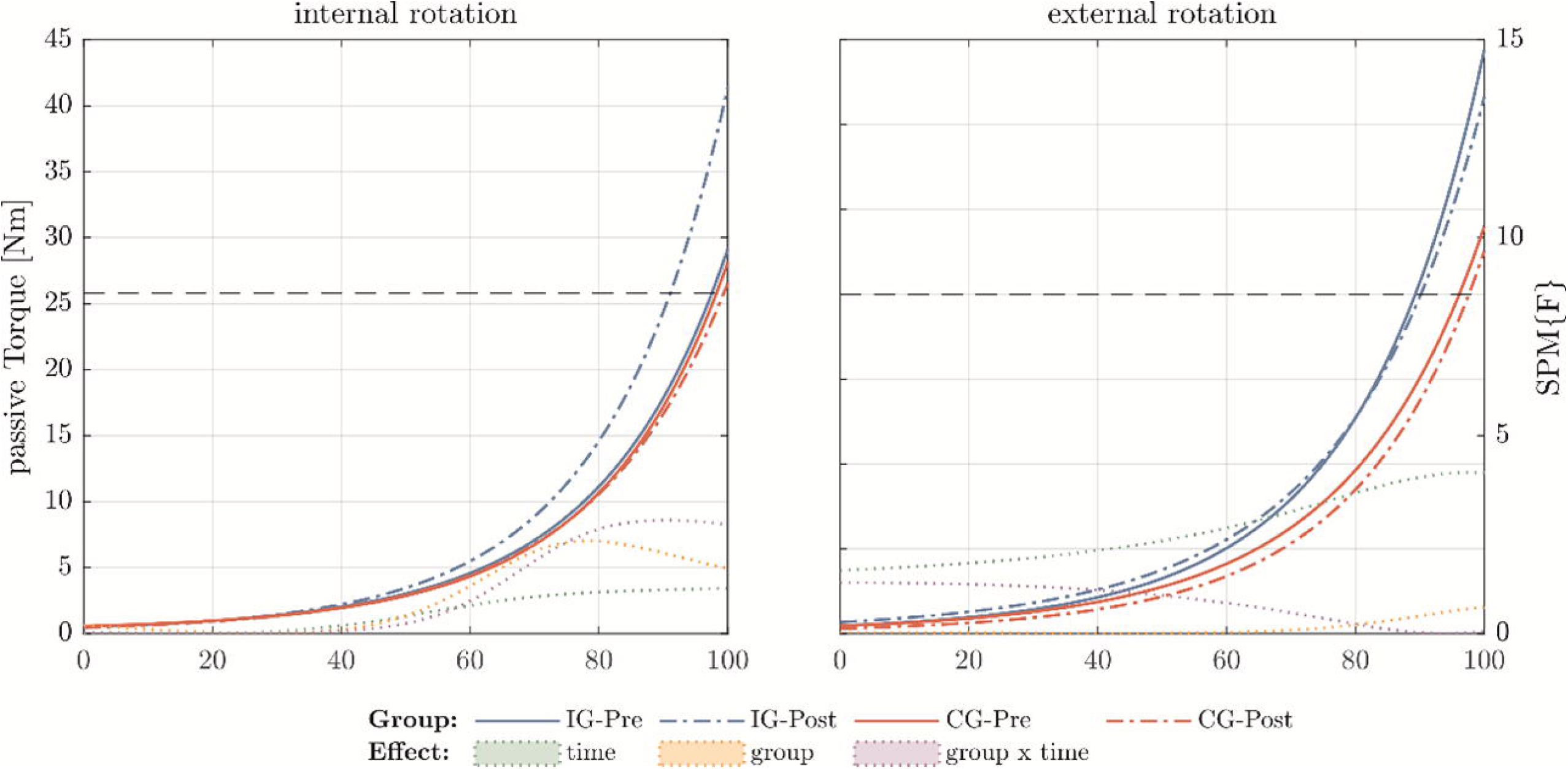
Passive torque-angle curves for the intervention group (IG, blue) and control group (CG, red), showing in the x-axis the range of motion until maximal stretch (100%). In the y-axis the F-value functions for the different effects time, group and time x group calculated using SPM1d. Values exceeding the critical F-value (dashed horizontal line) show significant differences for the different effects and are filled with the corresponding color.

### 3.2 Secondary outcome measures

Overall, for strength analysis the MANOVA did not reveal any effects for the internal rotator muscles. However, post-hoc analysis for the intervention group showed that eccentric and concentric strength increased in the 60°/s peak torque (Table 3). For flexibility measures, a two-way MANOVA (group x time) revealed no interaction effect for the passive stretching test for the internal rotator muscles (Wilks-Lambda = .932; F(2,22) = <1; p = .460; ηp^2^ = .068). In addition, post-hoc analysis showed positive pre-post changes in passive ROM for the intervention group (Table 3).

**Table 3:**
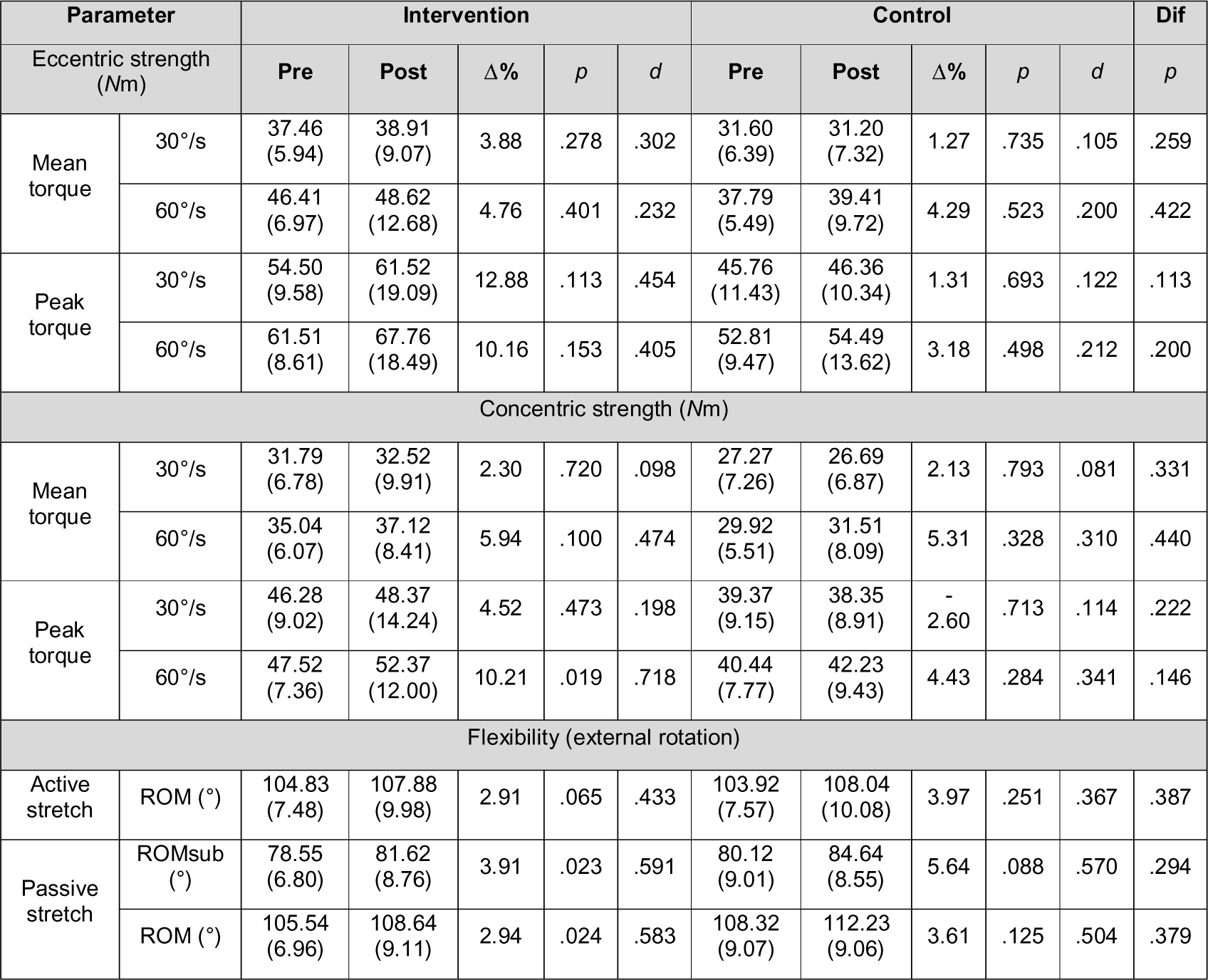
Secondary outcome measures post-hoc comparisons. Columns indicate the absolute pre and post-test values and in brackets the values for standard deviation for each group, percentage change (Δ%) and significant pre-post differences within groups (p), cohens d effect size (d), and in column “Dif” the statistical significance for between-group differences (normalized to body mass for strength). Nm, newton meter; ROMsub, submaximal range of motion; °, angle degree; °/s, degrees per second (movement velocity).

The internal rotator muscles showed group but not time differences in eccentric and concentric torque-angle relationship using 1SPMd analysis. Differences were found especially in the mid part of ROM and especially for the concentric condition (Figure 3).

**Figure 3:**
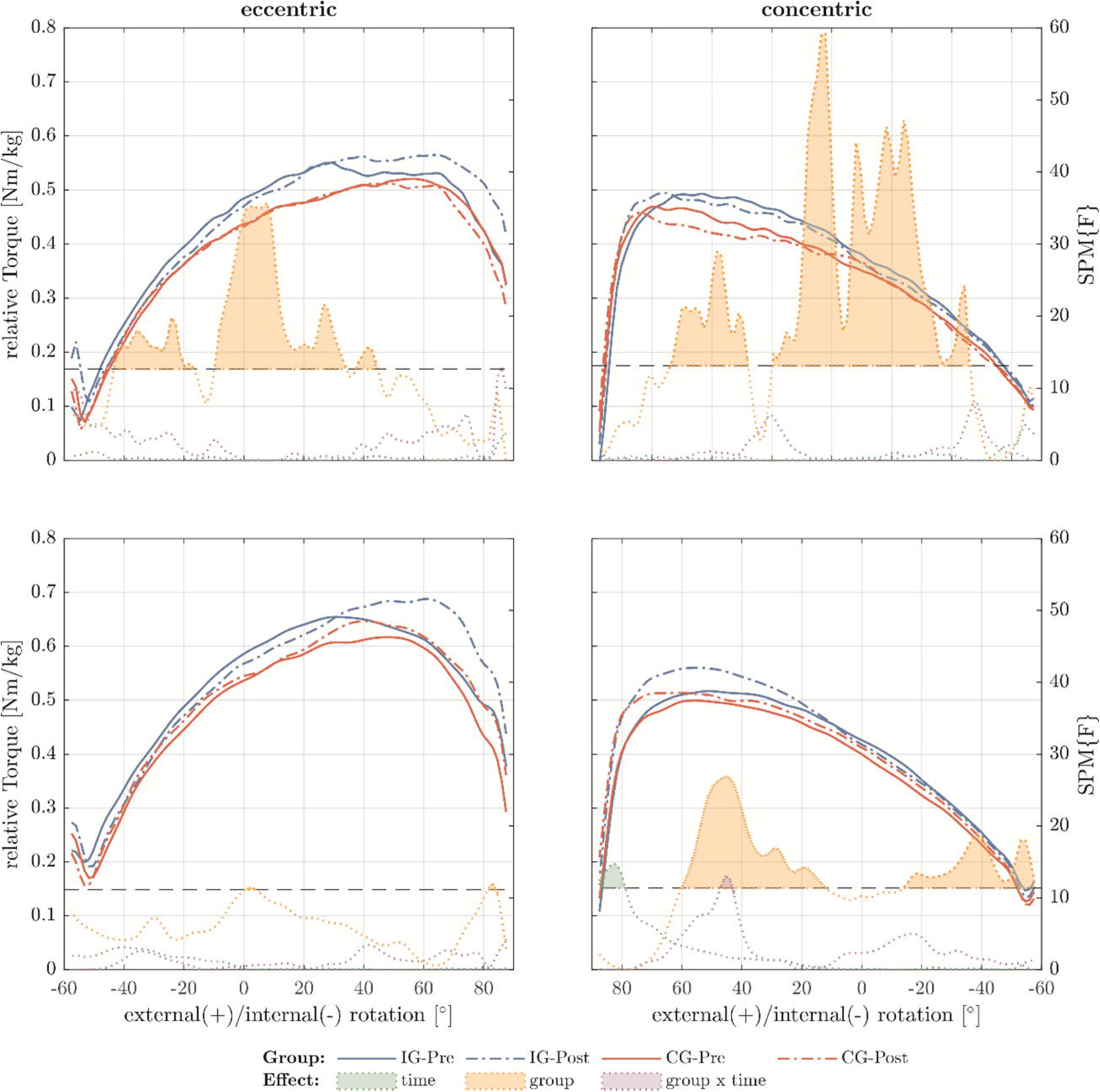
Relative torque-angle curves for 30°/s (top row) and 60°/s (bottom row) of the internal rotators for the intervention group (IG) and control group (CG) at the different time points of pre- and post-test. Additionally the F-value functions for the different effects time, group and time x group calculated using SPM1d. Values exceeding the critical F-value (dashed horizontal line) show significant differences for the different effects and are filled with the corresponding color.

## 4. Discussion

The aim of the study was to investigate functional and structural changes in the throwing shoulder of junior male handball players following a six week eccentric exercise compared to an active control group. We hypothesized that six weeks of 30°/s isokinetic eccentric resistance training for the external rotators would have positive effects on eccentric strength, flexibility, and supraspinatus and infraspinatus muscle FL and FV. Therefore, p-values for post-hoc comparisons were reported hypothesis dependent. However, in terms of statistical significance, the absolute values for eccentric strength increased independent of the movement speed by up to +15 %. Also, the supraspinatus fascicle showed +13 % lengthening, and 9 % change in the DTI metric FA which was significant different to the control group. The infraspinatus muscle FL increased for 8 % and the FA for 6 %. These findings were significantly different to the control group. No statistically significant changes where observed for flexibility measures and for the internal rotator measures.

First of all, the results presented were comparable to several eccentric-only intervention studies summarized by Vetter and colleagues.^19^ In this systematic review,^19^ a simple pooled analysis found that eccentric training for the lower limbs was more effective than conventional concentric training in changing more outcome parameters simultaneously, such as eccentric (+19 %), concentric (+9 %), passive ROM (+9 %), and FL (+10 %). Concentric strength training was found to be similarly effective in altering strength metrics, but did not alter other injury risk factors such as torque-angle relationship, passive ROM, or muscle FL.^19^ However, eccentric training can be interpreted as a beneficial approach to modifying multiple injury risk factors. Specifically, due to positive strength changes in the final ROM of each movement, for the throwing movement the important capability of energy absorption and energy transfer to concentric actions^6^ may be improved. These changes can also be interpreted as a reason for the comparably high decrease in complaints and injury rates explained following eccentric exercises that are regularly incorporated into athletic training routines.^16^

If comparing the present results to an eccentric training study for the shoulder, findings were partially similar.^23,30^ While Kim et al.^23^ showed that eccentric training is superior to concentric in terms of maintaining FL, Vetter and colleagues^30^ found a training effect for eccentric strength (24 %) and a 4 % decrease in ROM. Furthermore, Vetter and colleagues^30^ showed multiple positive effects on FL (+16 %) and FV (+19 %), but no changes occurred in the untrained internal rotators. This study recruited a sample of healthy, non-specific shoulder trained men for the same type of study design. While the results are comparable, the present study showed mostly smaller pre-post changes. This can be partly explained by a so-called plafond effect, as these recruited handball players show much higher absolute values in all functional and structural parameters. Another reason why these handball players showed slightly lower changes might be the starting handball season during the intervention phase. This may rise shoulder demands and produce concurring stimuli.^34^ In addition, the described in-season effects on shoulder function^34^ may also explain why external ROM increased in both groups in the present study, but was not statistically significant in the active control group.

An unchanged internal ROM following the eccentric training regime and an increase in passive torque-angle curve was also found by Vetter and colleagues.^30^ But in contrast to the mentioned preliminary study,^30^ the present sample may benefit from these results since an increase in torque-angle relationship in the final phase of eccentric contraction, an increase in passive torque and an unchanged ROM can be interpreted as a stabilizing and therefore significant preventive effect for the shoulder. Since Fieseler and colleagues^34^ describe a decrease in internal ROM during season, it confirms that an unchanged ROM may be interpreted as a positive result following the eccentric training during handball pre-season. The fact that the present results on joint flexibility stretching are based on a high-standardized stretching test, which has been evaluated in a preliminary study,^30^ shows that these results have to be interpreted with caution along with other studies.

In the present study, mDTI was used to demonstrate changes in muscle structural metrics following eccentric training. To the best of our knowledge, only Suskens and colleagues^25^ and a preliminary study for the shoulder used mDTI.^29^ FV is a metric first used in the study by Vetter and colleagues.^29^ As such, it is difficult to interpret and can be considered an exploratory metric. Interestingly in the present study, there were no statistically significant changes, but the supraspinatus muscle changed the most for the control group (12 %) than for the intervention group (8 %), in contrast to the preliminary study.^29^ These results may be plausible because positive changes in FL were reduced in the present study, which can be interpreted as correlating with muscle hypertrophy and changes in muscle thickness.^17^ Also interestingly, FA increased from pre to post and showed differences between the groups, which might indicate that the muscle fiber density and muscle state of health improved.^35^ Furthermore, the present study showed FL increases for 13 % and increases in FA for 6 %, whereas the preliminary study^30^ revealed no changes in FA and higher changes in FL (16 %). However, supraspinatus FL results can be compared to results from conventional 2D ultrasound measurements or cadaver dissections.^36,37^ While various research articles^36,37^ show very different FL for the supraspinatus muscle (2.8 to 8.3 cm), this study showed a mean FL of 3.7 to 4.2 cm. Very large heterogeneity in FL was also found for both investigated muscles in the present study. This is reasonable because the external rotator muscles can have very different volumes.^36^ Changes in FL may result from sarcomere lengthening or from sarcomerogenesis.^38^ However, we assume that muscle fiber remodeling after eccentric shoulder external rotator training is a functional correlate of sarcomere elongation based on previously reported changes in torque-angle curvatures.^39^

Several limitations have to be noted. First, the study design and sample size limited the power of the statistics. To the best of our knowledge, a comparable study by Kim and colleagues ^23^ recruited 13 subjects, while studies of the lower extremity have recruited more, suggesting group sizes of up to 16 subjects.^31^ However, we were unable to recruit more participants due to project limitations. Second, since the isokinetic training machine did not allow to define a target torque, the load magnitude was very heterogenous between subjects. Thus, subjects were told to finish their training session until they reached ± 10 % of an individual pre-defined workload based on the mean torque from the previous three sessions. Furthermore, the training load and compliance seemed to be playing position dependent. Third, four handball players seemed to have difficulties with the eccentric training and testing. These subjects did show very different torque-angle-relationships especially at the beginning and the end of the ROM during the strength tests. These players explained little discomfort in the shoulder when the experiment was finished. A fourth limitation might occur since this study appears to have primary research characteristics but also characteristics of a field experiment due to the implementation of the intervention in the regular athletics training regime. However, the use of an isokinetic dynamometer and MRI raised options of standardizations while in training the main goal was to complete twelve sessions within six weeks. Fifth, in terms of training exercise, the performed external rotation eccentric exercise seems to be less considered to force training-induced changes in muscle architecture compared to abduction exercises for the external rotator muscles.^23^ However, the idea was to focus on rotational movements based on the broadly known glenohumeral internal rotation deficit as a risk factor for injury.^40^ Last, besides the isokinetic submaximal passive flexibility test, which seems incomparable to other studies using goniometry,^19^ MRI showed limitations. A very heterogenous distribution in muscle volume within the recruited handball team led to incomplete representation of the external rotators using the largest available shoulder coil (XL). This has also been explained in a preliminary study.^30^ Following these difficulties, subjects showed different tract density after fiber tracking. Because of these difficulties, one subject had to be excluded due to artifacts in the supraspinatus muscle and only 15 subjects could be included in the analysis for the infraspinatus muscle due to a limited field of view in the MR images.

## 5. Conclusion

Eccentric strength training for the shoulder shows significant improvements in strength and muscle architecture, but not in flexibility measures, without decreasing shoulder joint flexibility in high trained junior handball players. The intervention group showed positive differences in eccentric strength and similar values for concentric strength compared to the active control group.

## 6. Perspectives

The strength of the present study is that it shows the advantage of an eccentric program due to multidirectional effects on eccentric torque, torque-angle relationship and muscle architecture. Furthermore, as the human shoulder joint is known to be a challenge for any imaging technique, the DTI results showed interesting insights into the shoulder tissue and its capability of adaptation following a training intervention. The implications are, first, that eccentric shoulder strength training could contribute not only to regular primary prevention and rehabilitation, but also to the performance development of the shoulder joint, and that future research projects should focus on different eccentric exercises and their multidirectional changes on the shoulder tissue.

## Data Availability

All data produced in the present study are available upon reasonable request to the authors.

## Acknowledgments

We thank all scientists and any assistive person involved in this study. A special thank goes to the handball club, its trainers and supportive stuff and to all handball players recruited for this study. Furthermore, a thanks goes to Simon Kiem who did support muscle segmentation. We also thank Frank Yeh for software support from DSI Studio. We confirm that all authors mentioned were involved in the present study and in the preparation of the manuscript.

## Authorship

**Sebastian Vetter:** Conceptualization; Funding Acquisition; Methodology; Investigation; Data Curation; Formal Analysis; Writing – Original Draft Preparation; Writing – Review & Editing;

**Maren Witt:** Conceptualization; Funding Acquisition; Methodology; Project Administration; Writing – Review & Editing

**Pierre Hepp:** Conceptualization; Resources; Writing – Review & Editing

**Axel Schleichardt:** Methodology; Investigation; Formal Analysis; Writing – Review & Editing

**Stefan Schleifenbaum:** Methodology; Writing – Review & Editing

**Christian Roth:** Investigation; Writing – Review & Editing

**Timm Denecke:** Resources; Writing – Review & Editing

**Jeanette Henkelmann:** Methodology; Investigation; Writing – Review & Editing

**Hans-Peter Köhler:** Conceptualization; Funding Acquisition; Methodology; Data Curation; Administration; Project Writing – Review & Editing

## Funding

This work was funded by the Federal Institute of Sports Science on behalf of the German Bundestag (grant number: ZMVI4-070601/22-23). The publication in a journal was funded by the publication fund of the University of Leipzig. The funders had no role in study design, data collection and analysis, decision to publish, or preparation of the manuscript.

## Conflict of interest statement

The authors declare that the research was carried out in the absence of any financial or non-financial interest that could be construed as a potential conflict of interest.

## Dataset

[dataset] Vetter S. (2023). Dataset_preliminary experimental study: Effect of isokinetic eccentric resistance training on strength, flexibility and muscle architecture. Figshare doi:10.6084/m9.figshare.24161040.v1

## Notes

### Competing Interest Statement

The authors have declared no competing interest.

### Clinical Trial

DRKS00028885

### Author Declarations

The Ethics Comittee at the Medical Faculty of the University of Leipzig raised no ehtical or sceintific objectives tot the study design presented. This study was granted 9 August, 2022, under the following ID: 362/21-ek

